# Is It Really Musculoskeletal Pain? Differential diagnosis and screening for referral in paediatric musculoskeletal disorders: a scoping review protocol

**DOI:** 10.1101/2021.11.23.21266780

**Authors:** A D’Aversa, F Bonetti, M Gallotti, F Maselli

## Abstract

**Introduction:** Musculoskeletal disorders are common in children and adolescents. They could have a broad spectrum of causes, which the most are self-limiting and usually do not need a referral to specialistic care. It is mandatory for the healthcare professionals working primary care settings to be able to understand when to perform a “wait and see” approach or when further investigations are needed. This study aims to identify signs and symptoms reliable to serious or life-threatening conditions that could be detected during the screening process and the physical examination of children and adolescent with common musculoskeletal disorders by a healthcare professional in direct access clinical setting.

**Materials and Methods:** The scoping review (ScR) will be performed following the six-stage methodology suggested by Arksey and O’Malley and the extensions to the original framework that are recommended by the Joanna Briggs Institute. Fourteen electronic databases will be investigated since their inception. The Preferred Reporting Items for Systematic reviews and Meta-Analyses extension for Scoping Reviews (PRISMA-ScR) checklist will be used to meet the required standards of quality and reproducibility. Review tools will be used for the quality assessment of the included studies. This protocol received input from all authors who have expertise in research methodology and paediatric and musculoskeletal disorders.

**Ethics and Dissemination:** The results of this ScR will be relevant given that screening for referral in paediatric musculoskeletal disorders has not been addressed in the literature properly yet. Dissemination will include submission to peer-reviewed journal. Ethical approval is not required for scoping reviews.

**PROTOCOL INFORMATION:** Anticipated or actual start date: January 2021

Anticipated completion date: December 2021

## INTRODUCTION

Musculoskeletal disorders are one of the most common causes of disability for the global population. [1] Particularly, musculoskeletal disorders (MD) have already been studied extensively among adults but they are very frequent even among children and adolescents, affecting up to 30% of individuals. [2,3] There is a broad spectrum of causes, most of which are self-limiting, frequently traumatic, and often do not need a referral to specialist care. [4] Unfortunately, these causes can also include potentially life-threatening conditions, or could be an early sign of future chronic diseases, both of which should be properly detected and managed. [5,6] To properly frame each MD in childhood patients, an appropriate triage based on careful clinical assessment, an efficient use of investigations, a deep knowledge of common and medically significant conditions and awareness of local referral pathways are mandatory. [7,8]

Failure to recognize these conditions may result in treatment delay, significant morbidity, future health problems, excessive use of health care resources and redundant, sometimes invasive diagnostic tests and overmedicalization. [9] Musculoskeletal pain that has been interpreted as benign or idiopathic, respectively, as well as injuries that remain unexplained, can jeopardize a child’s welfare by causing chronic pain syndromes with substantial psychological and physical impairment. Moreover, the primary care physician must recognize when further intervention, is needing using the minimum amount of testing to make an appropriate, prompt diagnosis, rather than reassurance, follow-up and subspecialty referral. However, considering the expanding spectrum of diagnostic options, this can be a challenge today. [10]

### Aims

This study aims to identify signs and symptoms reliable of serious or life-threatening conditions, which mimic non-specific musculoskeletal disorders during the screening process and the physical examination of children and adolescent by healthcare professionals in direct access clinical setting. It will be a scoping review of peer-reviewed empirical studies relating to definitions of RFs in pediatric musculoskeletal disorders. The secondary target will be to provide a critical overview of current empirical literature to aid identification of future research trends in screening for referral.

## MATERIAL AND METHODS

The approach to be taken to systematically review the knowledge based on our topic of interest was determined by considering the purpose of this review and the nature of the data searched. A detailed and sensible search string has been created to achieve a broad and thorough examination of the available literature related to the early detection of life-threatening conditions, evaluation of RFs in pediatric musculoskeletal disorders and the latest guidelines for screening for referral approach. For the purposes of this scoping review, the source of information can include any existing literature.

### Protocol design

The protocol design is guided by Arksey and O’Malley’s methodological framework to conduct scoping review, which includes six steps [11], and the extensions to the original framework recommended by the Joanna Briggs Institute (JBI). [12] The Preferred Reporting Items for Systematic reviews and Meta-Analyses extension for Scoping Reviews (PRISMA-ScR) checklist will be used to ensure that the scoping review follows necessary qualitative standards of practice and reproducibility. [11] We will follow the acronym of Population, Concept and Context (PCC) proposed by The Joanna Briggs Institute to describe the elements of the inclusion criteria.

#### Population

The studies included will report data about 0 to 18 years old individuals of both sex that must have at least one condition that mimics any musculoskeletal disorder.

#### Concept

The main concept of this review will be to list any sign or symptom reliable to serious or life-threatening conditions, which mimic musculoskeletal disorders during the screening process and the physical examination of children and adolescent by a healthcare professional in direct access clinical setting.

#### Context

Literature concerning pediatric musculoskeletal disorders will be reviewed. Experimental and clinical settings will be considered. However, we will focus mainly on the studies which relied on measures, methodologies, and/or tests that are administrable in clinical practice.

First, to identify research studies, a systematic search of the peer-reviewed literature using medical subject headings (MeSH) terms, keywords and database categorization will be conducted. This iterative process will involve broad search terms and tips, including truncations and synonymous of “differential diagnosis”, “red flags”, “musculoskeletal disorders”, “musculoskeletal disease”, “children” and “adolescent” in Medline (PubMed), Embase, Cochrane Library includes the Cochrane Central Register for Controlled Trials, CINAHL complete, Scopus, Web of Science, Pedro, Uptodate, Living OVerview of Evidence (LOVE), Epistemonikos, Diagnostic Test Accuracy database (DiTA), TRIP Medical Database, Google Scholar, OpenGrey. The grey literature will be screened using Google database and the search will be conducted using a modified version of the final search strategy.

All reference lists of included studied will be searched manually to identify any additional studies that may be relevant for this review, then these articles will be stored and screened using Rayyan QCRI software. The screening of the identified studies will be completed in two steps. Studies available in English, Italian, German, French or Spanish language will be included. No date limitations will be imposed for the search strategy.

A data collection form will be developed and reviewed by authors to confirm study relevance and to extract study characteristics.

Due to the potentially large volume, breadth and heterogeneity of material included in a scoping review, it is not possible to predetermine the optimal method to collect and reporting the results. Reporting methods will be accurately documented for readers to assess for any potential reporting bias. [13] The exact subset will be determined during the research process with a rationale for conducting a critical appraisal. [14-16]

## DISCUSSION

The studiers of differential diagnosis process and the subsequent screening for referral have been very thorough in the adult population in last years, [17-20] but a little has been dedicated to study the pediatric patients, therefore we aim to review scientific literature published on the pediatric musculoskeletal disorders.

This study aims to identify signs and symptoms reliable of serious or life-threatening conditions, which mimic non-specific musculoskeletal disorders during the screening for referral process through the history taking and physical examination of children and adolescent by a healthcare professional in direct access clinical settings. To the best knowledge of the authors this is the first study that investigates these topics. Through the publication of this protocol we intend to define and to outline the step-by-step procedure to conduct the scoping review in order to limit the occurrence of reporting bias. It provides a critical overview of current empirical literature to aid identification of future research trends in screening for referral.

## Data Availability

All data produced in the present study are available upon reasonable request to the authors

## DISSEMINATION PLANS AND ETHICS

No ethical approval was required for this literature-based study.

The dissemination process will include the publishing of the results and summary of the review and presentation of the study in national and international conferences and networks on musculoskeletal disorders.

### Funding

No financial or material support of any kind was received for the work described in this article.

### Contributors

All authors designed the protocol, reviewed the manuscript, approved the final version and participated in the six steps.

**Table I.** Eligibility criteria for included and excluded studies.

| *Inclusion criteria* | *Exclusion criteria* |
| --- | --- |
| Studies investigating signs and symptoms (red flags) mimicking a musculoskeletal disorder in paediatric age. | Studies investigating in paediatric age that not mimicking musculoskeletal disorders. |
| Language: full text in English, Italian, German, French, Spanish | Language: full text not in English, Italian, German, French, Spanish |
| Setting: experimental or clinical (with a particular focus on studies reporting methodologies or test/measures feasible to translate into clinical practice). |  |
| Participants: studies on human, children or adolescent (0 to 18 years old), male and female, affected by musculoskeletal disorders or at least one condition that mimics musculoskeletal disorders. | Participants: studies on humans, over 18 years old, affected by condition that not mimics musculoskeletal disorders. |
| Study design:<br>*Primary research studies*<br>Quantitative design:<br>- Experimental designs (randomized controlled trials, controlled clinical trials, proceedings, conferences abstracts)<br>- Observational designs (descriptive studies, surveys, cohort studies, cross-sectional studies, observer-reported or patient-reported outcome studies, case studies and case series, proceedings, conference abstracts, ecological studies)<br>*Secondary research studies*<br>- of quantitative studies: systematic review with or without meta-analysis, observational<br>- of qualitative studies: meta-summary, meta-synthesis<br>- narrative review<br>- editorials<br>- commentaries or expert opinion articles<br>- point of view<br>- book reviews or chapters<br>- study protocol | Study design:<br>- letters to editors<br>- correspondences or replies to letters |
| Outcomes:<br>- healthcare practitioner<br>- setting (primary care, hospital, physiotherapist clinic...)<br>- validated outcome measures as RFs' cut-off<br>- risk factors (for RFs) |  |

